# A multimodal AI biomarker PATH-ORACLE improves prediction of recurrence in stage I lung adenocarcinoma

**DOI:** 10.64898/2026.01.28.26344973

**Authors:** Oz Kilim, Orsolya Pipek, Zsofia Sztupinszki, Miklos Diossy, Aurel Prosz, Cristina Naceur-Lombardelli, Selvaraju Veeriah, David Moore, Mariam Jamal-Hanjani, Allan Hackshaw, Janos Fillinger, Judit Moldvay, Istvan Csabai, Charles Swanton, Zoltan Szallasi

**Affiliations:** Department of Physics of Complex Systems, Eötvös Loránd University, Budapest, Hungary; Computational Health Informatics Program, Boston Children’s Hospital, Harvard Medical School, Boston, MA, USA; Danish Cancer Institute, Copenhagen, Denmark; Department of Pathology, National Koranyi Institute of Pulmonology, Budapest, Hungary; 1st Department of Pulmonology, National Koranyi Institute of Pulmonology, Budapest, Hungary; Pulmonology Clinic, University of Szeged, Albert Szent-Gyorgyi Medical School, Szeged, Hungary; Cancer Evolution and Genome Instability Laboratory, The Francis Crick Institute, London, UK; Cancer Research UK Lung Cancer Centre of Excellence, University College London Cancer Institute, London, UK; Department of Bioinformatics, Semmelweis University, Budapest, Hungary; Translational Oncopulmonology Research Group, Institute of Molecular Life Sciences, HUN-REN Research Centre for Natural Sciences, Budapest, Hungary; Cancer Metastasis Laboratory, University College London Cancer Institute, London, UK; Department of Medical Oncology, University College London Hospitals, London, UK; Cancer Research UK & UCL Cancer Trials Centre, London, UK; Department of Cellular Pathology, University College London Hospitals NHS Foundation Trust, London, UK; Cancer Research Institute, University College London Cancer Institute, London, UK

**Author notes:** Correspondence should be sent to: Zoltan Szallasi: Computational Health Informatics Program (CHIP) Boston Children’s Hospital, Harvard Medical School, 300 Longwood Ave., Boston Massachusetts, USA, 02215, Istvan Csabai: Eötvös Loránd University, Department of Physics of Complex Systems, H-1117 Budapest, Hungary, Pázmány Péter sétány 1/A (5-th floor, office: 5.64), Charles Swanton: Cancer Evolution and Genome Instability Laboratory, The Francis Crick Institute, 1 Midland Road, London, UK, NW1 1AT; Cancer Research UK Lung Cancer Centre of Excellence, University College London Cancer Institute, 72 Huntley Street, London, UK, WC1E 6DD; Department of Medical Oncology, University College London Hospitals, 235 Euston Road, London, UK, NW1 2BU.

**Keywords:** Early-stage Lung adenocarcinoma, Reccurance prediction, Multimodal deep learning

## Abstract

The standard treatment for stage I lung adenocarcinoma is surgical resection, in most cases without additional systemic adjuvant treatment. A significant proportion of stage I cases recur with a less than 50% 5-year survival rate. There are clinical data suggesting that adjuvant treatment may improve survival in such recurrent cases. However, previously evaluated predictors such as the IASLC grading system from histological sections and transcriptomic profiles have not been sufficiently accurate and consistent for risk stratification and to guide therapeutic interventions. We hypothesized that these previously investigated diverse diagnostic measurements carry complementary information that may provide higher prognostic power when combined. Here we describe a multimodal deep learning method, PATH-ORACLE. This biomarker is built on top of the prospectively validated transcriptomic-based ORACLE score with the addition of routine histological sections processed by pre-trained foundation models. PATH-ORACLE predicts recurrence with an accuracy of over 85% in two independent cohorts. Given further validation this predictor could be used to prioritize stage IB patients for adjuvant chemotherapy in a more consistent fashion. Furthermore, for stage IA cases, PATH-ORACLE, combined with liquid biopsy-based monitoring may help identify high-risk patients suitable for adjuvant targeted therapy.

**Highlights:**

- Multimodal AI model (PATH-ORACLE) integrates histology and transcriptomics to predict stage I LUAD recurrence
- PATH-ORACLE outperforms IASLC grading and transcriptomic or image-based models alone
- Model achieves >85% recurrence prediction accuracy across independent international cohorts
- PATH-ORACLE refines risk stratification within both stage IA and IB lung adenocarcinoma
- Biomarker may guide adjuvant therapy selection and surveillance in early-stage disease

## 1 Introduction

Surgical resection with mediastinal lymphadenectomy is the standard treatment for patients with stage I lung adenocarcinoma (LUAD). For stage IA patients, despite the 10-20% 5-year recurrence rate, no further systemic treatment is currently recommended (1–3). The 5-year recurrence rate for stage IB patients is even higher, up to 35%, but adjuvant chemotherapy is still optional and recommended only for patients that are considered to be high risk based on histologic risk factors such as tumor size, tumor grade, lymphovascular invasion, and visceral pleural invasion (4). For patients with resected stage IB adenocarcinoma and epidermal growth factor receptor (EGFR) exon 19 deletion or exon 21 L858R point mutations, adjuvant targeted therapy with osimertinib is recommended (5). Lung cancer relapse is associated with significantly worse clinical outcome and reduced overall survival. An accurate predictor of relapse will allow the close monitoring of high-risk patients, by e.g. the genomic analysis of liquid biopsies, and systemic intervention could be initiated with likely long-term clinical benefit (6).

Several types of diagnostic data modalities have been evaluated for predicting recurrence in early-stage LUAD. Pathological classification, such as the 3-tiered International Association for the Study of Lung Cancer (IASLC) grading system (7), is informative of clinical outcome in early stage I cases (8). The interobserver variability of estimates of the proportion of the various histological subtypes within a given tumor was recently circumvented by an automated image analysis algorithm (9), further confirming the utility of histological classification for predicting recurrence in early stage LUAD.

Gene expression-based predictors, such as ORACLE (10,11), and RiskReveal (AIM-HIGH) (12) have a clinically validated performance of predicting outcome when several stages of LUAD are evaluated together, but can be improved for stage I cases. Mutational profiling based on next generation sequencing was also evaluated, with limited utility to predict relapse in stage I LUAD (13). Importantly, while the power of these methods to predict recurrence of stage I LUAD is significant, their accuracy (usually in the range of AUC 0.6-0.65) does not reach the level that would determine changes in therapeutic interventions.

If the diagnostic information of these different diagnostic methods is complementary, then it is reasonable to assume that their combination may yield a more accurate predictor of clinical relapse. Previously, we successfully tested this assumption in high-grade serous ovarian carcinoma, where the combination of histological images with proteomics yielded an independently validated predictor of response to first-line platinum-based therapy (14).

Traditionally, combining histological images with gene expression profiles or genomics is not a trivial task, but the emergence of multi-modal deep learning has provided a computationally feasible strategy of combining such disparate diagnostic features.

Here, we performed a multimodal deep learning training of a predictor of recurrence of early-stage LUAD on four independent cohorts covering 710 patients from three geographic regions, where matched H&E histological images and gene expression measurements were both available. We found that the multimodal deep learning combination we term **PATH-ORACLE** provides a more accurate predictor of relapse in stage I LUAD compared to the transcriptomics or histology based prediction alone.

## 2 Results

### 2.1 Improving prognostic performance for stage IA/IB LUAD in unimodal settings

#### 2.1.1 Deep learning-based classification of H&E whole slide images is a more accurate predictor of recurrence than the IASLC grading system

Histological features of LUAD are associated with patient prognosis (8,15). Currently, the updated IASLC grading system can be used for risk stratification from H&E whole slide images (WSI) (7) **(Fig 1.a)**. This grading system is based on the 5th edition of the World Health Organization (WHO) classification of five main architectural patterns (**Fig 1.b**) in invasive non-mucinous pulmonary adenocarcinoma: lepidic, acinar, papillary, micropapillary, and solid (16). This system is implemented by pathologists manually, visually estimating the percentage of each pattern in 5-10% increments.

**Figure 1.**
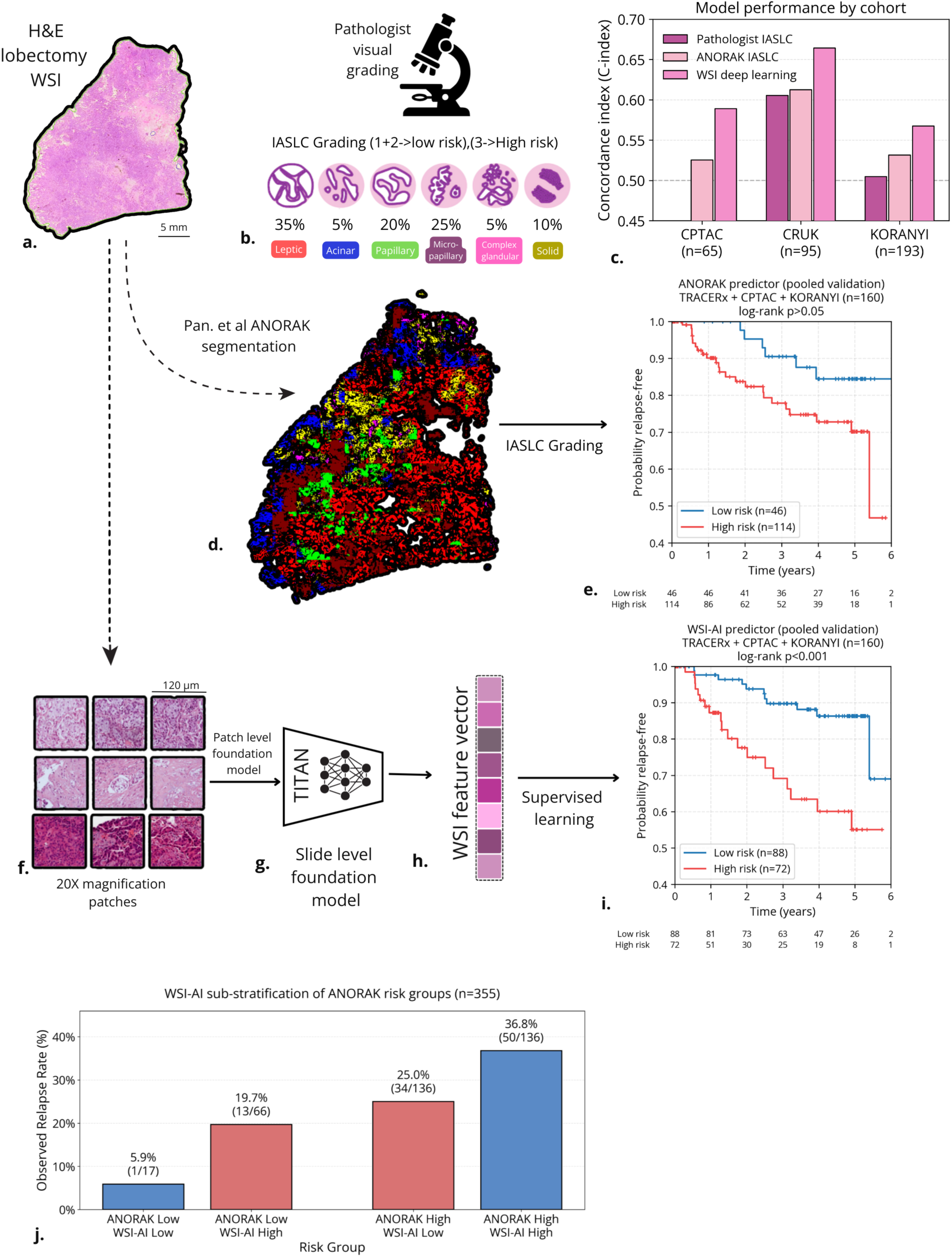
H&E WSI analyses. **a**. H&E WSIs are gigapixel images scanned at 20X magnification. **b**. Slide classification is performed by pathologists through visual assessment in 5% increments following IASLC morphologic guidelines. **c**. Comparing Pathologist based grading with ANORAK and our WSI based deep learning model for three validation cohorts. Performance comparison C-index for RFI for CPTAC, TRACERx, and KORANYI cohorts. The WSI level deep learning predictor outperforms manual and ANORAK-based grading. **d**. ANORAK (9) slide level segmentation into histotypes; Leptic, Acinar, Papillary, Micropapillary, Complex glandular, and Solid. **e**. IASLC grading (1+2 low risk, 3 high risk) from ANORAK based histotypes stratifying collated validation cohorts by risk. **f**. Segmenting tissue from background and patching WSIs into smaller RGB images for embeddings. **g**. Whole slide level embedding of patch level Conch1.5 (17) embeddings with TITAN(18). **h**. The 783-dimensional feature vector representing the WSI used for downstream prediction of relapse risk. **i**. The WSI foundation model-based model significantly stratifying collated validation cohorts by risk. **j**. WSI AI-based predictor subdivides ANORAK-based grading groups revealing high risk cases ANORAK defines as low risk and low risk cases ANORAK defines as high risk.

To establish a baseline, we evaluated the prognostic performance of pathologist-assigned IASLC grades across our validation cohorts. Single pathologist assessments showed consistently weak associations with relapse-free survival across two validation cohorts where pathologist assessments were available **(Fig 1.c)**: TRACERx (C-index = 0.61, 95% CI [0.49–0.72], log-rank p = 0.067, HR = 2.40 [0.91–6.32]) and KORANYI (C-index = 0.50, 95% CI [0.45–0.56], log-rank p = 0.881, HR = 1.04 [0.60–1.81]). The CPTAC cohort was not analyzed as it was underpowered for this analysis since only 30% of histopathology slides were graded by a pathologist. These results highlight the prognostic yet still limited predictive power of IASLC grading based on single pathologists’ evaluation in stage IA/IB disease. These findings are consistent with large scale (n= 1443) evaluations of IASLC grading (8).

The pyrAmid pooliNg crOss stReam Attention networK (ANORAK) addresses key limitations in manual IASLC grading, particularly interobserver variability and inconsistent quantification of growth patterns (9). ANORAK uses pixel-wise segmentation for automated, objective IASLC grading, reducing subjectivity in differentiating lepidic, papillary, and acinar patterns (**Fig 1.d**).

We applied ANORAK to generate automated IASLC grades across the three validation cohorts CPTAC, TRACERx, and KORANYI. ANORAK-derived grades showed modestly improved prognostic stratification in stage IA/IB LUAD compared to the single pathologist assessment. TRACERx (C-index = 0.61, 95% CI [0.50–0.70], log-rank p = 0.102, HR = 2.20 [0.84–5.78]), CPTAC (C-index = 0.53, 95% CI [0.50–0.56], log-rank p = 0.490), and KORANYI (C-index = 0.53, 95% CI [0.48–0.58], log-rank p = 0.132, HR = 1.75 [0.84–3.67]) (**Fig 1.c**). Pooling all validation cases, ANORAK significantly stratifies high and low risk group cases (log rank p<0.05) **(Fig 1.e).** While ANORAK automates the pathologist-defined workflow, its reliance on previously defined histopathological features limits its ability to leverage any other complex morphological patterns that might exist with potential prognostic relevance beyond the IASLC framework.

To check if such patterns exist, we trained a WSI foundation model-based relapse predictor on the stage IA/IB sub-cohort of TCGA-LUAD (n=272). This data driven approach enables automatic discovery of relapse related features. Whole slide images were first patched and embedded at 0.25 µm^2^ resolution using Conch1.5 (17) (**Fig 1.f**). These patch-level embeddings were then aggregated into slide-level representations using TITAN (18) **(Fig 1.g)** (see Methods), yielding a fixed 768-dimensional vector for each slide **(Fig 1.h)**. We applied regularized Cox regression to learn relapse risk directly from censored and uncensored follow-up data in the TCGA-LUAD cohort. We validated the model across three independent cohorts (TRACERx, CPTAC, and KORANYI) using a fixed predefined model and evaluation pipeline.

The WSI foundation model-based predictor significantly outperformed both pathologist-assigned and ANORAK-derived IASLC grades across three independent validation cohorts (**Fig 1.c**). External validation demonstrates robust generalization. For a 1-year cutoff AUC the validation cohorts reached accuracy of CPTAC = 0.659 (n=48), TRACERx = 0.756 (n=92), and KORANYI = 0.634 (n=195), For 2-year cutoff AUC scores, results were 0.75 in CPTAC (n=30), 0.75 in TRACERx (n=88), and 0.66 in the KORANYI blind test cohort (n=193) (**Fig 1.c**). The C-index values were CPTAC = 0.60 (n=68), TRACERx = 0.67 (n=112), and KORANYI = 0.58 (n=272) (95% CI [0.37–0.76], [0.53–0.79], and [0.50–0.64], respectively). These results demonstrate that the WSI foundation model captures relapse-specific morphological signatures that generalize across institutions, scanners, and patient populations. When pooling all validation cases, this deep learning-based model significantly stratifies high and low risk group cases (log rank p<0.001) **(Fig 1.i).**

In direct comparisons, the WSI foundation model-based predictor achieved superior prognostic stratification compared to ANORAK IASLC grading (C-index Δ=+0.10; 95% CI [0.03–0.18]; bootstrap p=0.011).

The WSI predictor reveals prognostic heterogeneity within IASLC grades. To explicitly investigate whether the WSI model captures information beyond IASLC grading, we stratified patients by their ANORAK based IASLC grade and further subdivided them by WSI-deep learning based predicted risk. The WSI predictor identified high-risk cases within the IASLC Grade 1+2 low-risk group and lower-risk cases within the IASLC Grade 3 high-risk group (**Fig 1.j**). This further suggests that the foundation model-based approach captures fine-grained morphological patterns not encoded in the IASLC framework.

#### 2.1.2 RNA seq analysis – A LUAD Stage I relapse specific ORACLE signature

Transcriptomic markers could improve diagnostic precision in LUAD by identifying molecular subtypes associated with relapse risk and mortality (11,19). These profiles capture gene-expression programs and intracellular signaling states that may not be fully reflected in histopathological images and may contain complementary prognostic information.

The most promising approach is the Outcome Risk Associated Clonal Lung Expression (ORACLE) that additionally takes into spatial gene expression variability within a tumour (10). This is a prospectively validated 23-gene transcriptomic biomarker trained on TCGA stage I-III cases for overall survival (OS) (11). It stratifies non-small cell lung cancer patients into high or low-risk survival categories.

We ran a multi cohort validation on the ORACLE signature and 7 other transcriptomic signatures published in the last 5 years (10,20–26) to understand how robust these are for stage I cases relapse prediction specifically (**Fig 2.a**). Training on the combined gene set from the three best performing gene signatures from independent studies [ORACLE(11), Zhao(21), and Carr(20)] improved prognostic performance. Sparse Cox regression identified 8 genes [LRP12, ANLN, PNP, UPK18, XBP1, PEBP4, EDN3] that provided a strong signal for stage IA/IB relapse risk (**Fig 2.b**).

**Figure 2.**
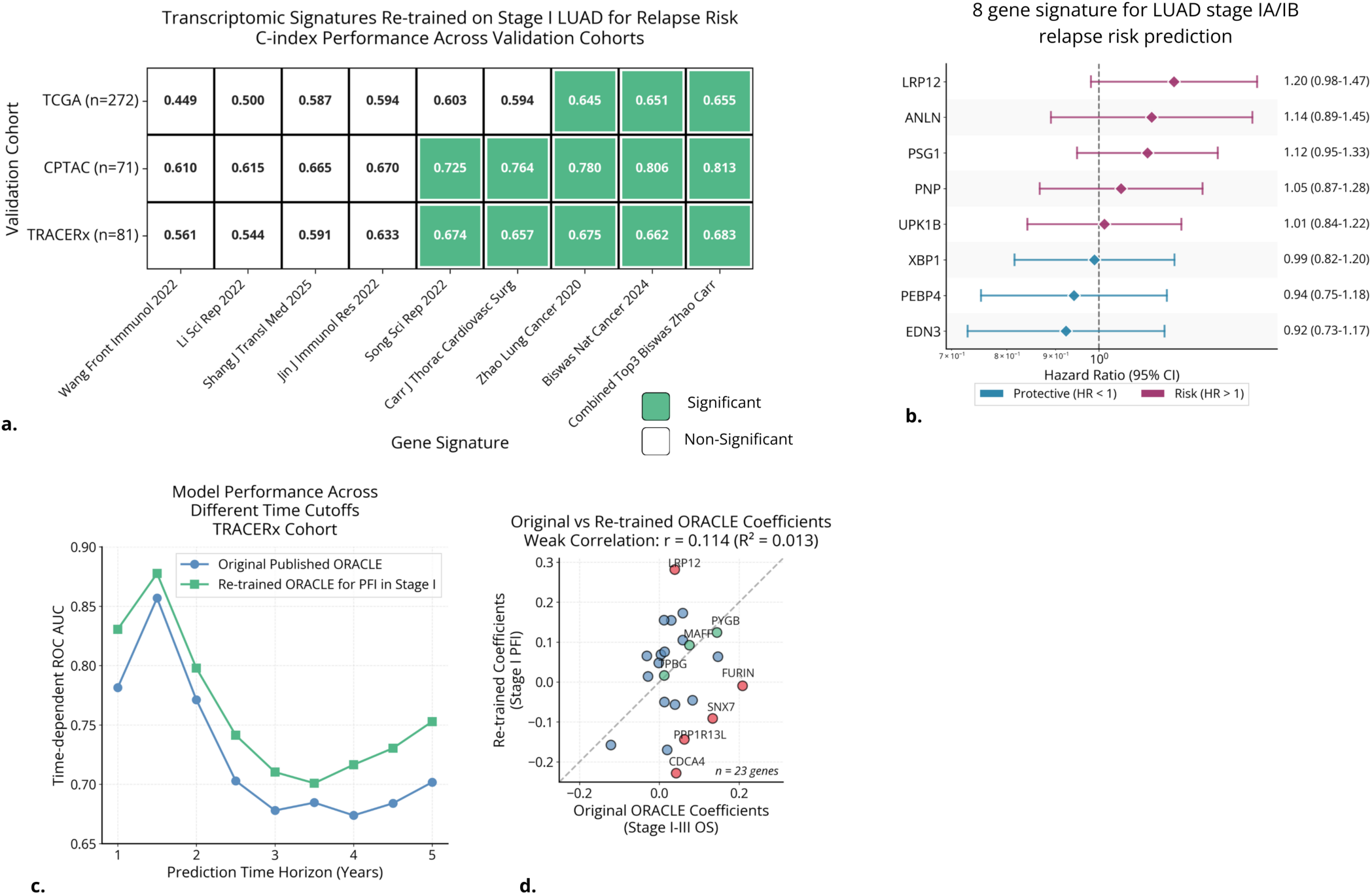
RNA-Seq analysis. **a**. C-index for various transcriptomics gene signatures across TCGA-LUAD 5-fold cross-validation, CPTAC external validation, and TRACERx external validation. **b**. Set of discovered genes combining top three signature gene set (Carr, Zhao, Biswas) and training for RFI Stage I cases only. **c**. ORACLE (11) gene signature compared to the TCGA-LUAD RFI stage I only re-trained 23 gene signature. **d**. Correlation between original ORACLE signature coefficients and re-trained coefficients.

We found we could enhance robustness for stage IA/IB prediction, by re-training gene signatures using RNA-Seq profiles for stage IA/IB cases explicitly for relapse instead of OS. ORACLE performance improved when retrained specifically with stage IA/IB cohorts for relapse risk cases (C-index TRACERx=0.66, CPTAC=0.81, TCGA 5-fold Cross validation (CV)=0.66) compared to the OS stage I-III trained ORACLE (C-index TRACERx=0.64, CPTAC=0.70, TCGA 5-fold Cross validation (CV)=0.62) (**Fig 2.c**). This suggests that retraining on the target IA/IB subset of patient data allows the model to capture stage-specific biology obscured when grouping Stages I–III. The models were most predictive of relapse within 6 months to 2 years. We observed improved C-index scores of 5-10% across all time-until-relapse cutoffs over the original ORACLE signature (**Fig 2.c**).

Interestingly, the learned coefficients on re-training showed weak to no correlation (Pearson r = 0.1143, p = 6.04e-01, Spearman ρ = 0.0227, p = 9.18e-01) with those from the original Stages I–III, OS training (**Fig 2.d**). To rule out the chance of technical artifacts from stochastic instability in training, we retrained the RNA-Seq model using 5 seeds and 10 CV folds. Fold-to-fold and seed-to-seed correlations were high (Pearson r ≈ 0.75–0.95) suggesting no such artifacts.

Comparing the original and stage I re-trained ORACLE, the most inconsistent genes included CDCA4, LRP12, SNX7, FURIN, and PPP1R13L, while the most consistent genes included TPBG, MAFF, and PYGB.

To extend these findings, we investigated whether this newly identified Stage I relapse-specific transcriptomic signature could be integrated with WSI-based data to capture orthogonal signals and improve risk prediction.

### 2.2 PATH-ORACLE: Multimodal deep learning combining histological images and gene expression profiles outperforms previous predictors of relapse in multiple, independent cohorts

We constructed a novel Multimodal deep learning model PATH-ORACLE, by concatenating embeddings, then combining WSI features discovered in the WSI-based analysis and the RNA-Seq signatures discovered in the RNA-Seq only analysis section. We found that histopathological and transcriptomic features are additive i.e. the prediction accuracy of PATH-ORACLE is larger than its constituent modalities **(Fig 3.a)**. For the cases with both WSIs and RNA-Seq data in the validation sets, model performance was: CPTAC, AUCs were 0.86 [0.69–0.98], p=.002 (n=43) at 1 year; 0.86 [0.67–0.99], p=0.0021 (n=24) at 3 years; and 0.97 [0.86–1.00], p=0.005 (n=15; 12 events, 3 nonevents) at 5 years; TRACERx, AUCs were 0.84 [0.64–0.99], p=0.0028 (n=65) at 1 year; 0.71 [0.54–0.86], p=0.0065 (n=66) at 3 years; and 0.78 [0.61–0.91], p=0.0014 (n=39) at 5 years **(Fig 3.b)**. Together, both cohorts show strong early discrimination, with sustained long-term performance in CPTAC and moderate but significant long-term discrimination in the TRACERx cohort.

The multimodal model consistently improves over WSI alone. Pooled across TRACERx + CPTAC the C-index is 0.70 for PATH-ORACLE vs 0.63 for the WSI model. On CPTAC only, the gain is statistically significant (C=0.72 vs 0.58; bootstrap p=0.03), and on TRACERx, the model showed improvement as well, although to a lesser extent (C=0.72 vs 0.64) p=0.26. Importantly, using a single cutoff (k=24) for the risk scores predicted by PATH-ORACLE, patients can be efficiently stratified into high- and low-risk groups for relapse across multiple validation cohorts (TRACERx P=0.0004, HR=4.99, [CI 1.87,13.28] CPTAC P=0.0003, HR=6.90 [2.02-23.63]). PATH-ORACLE was further compared against established risk factors of relapse and significantly outperformed them (**Fig 3.c**.), highlighting potential clinical utility in these findings.

**Figure 3.**
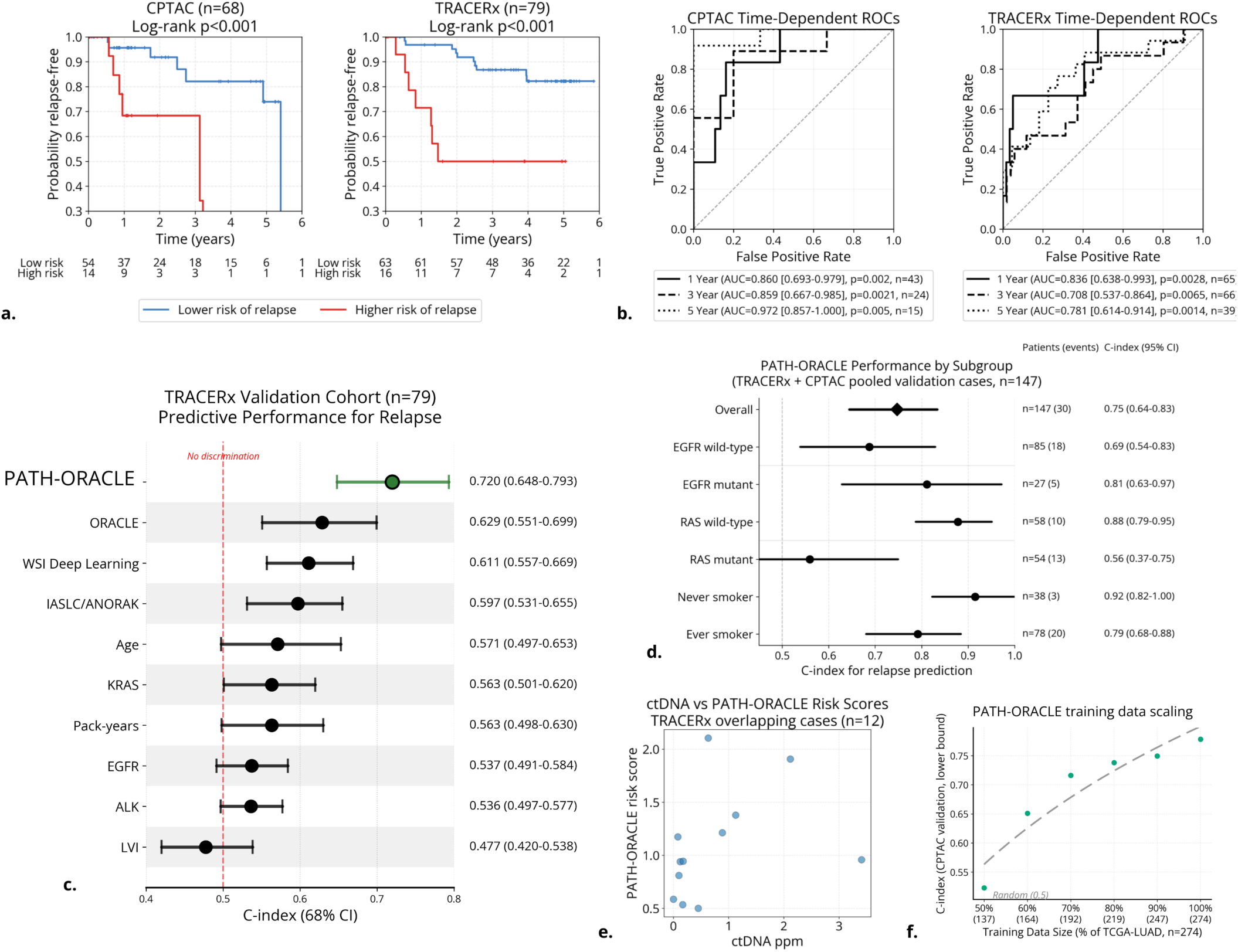
Multimodal analysis. **a**. KM plots evaluating prognostic performance of PATH-ORACLE on CPTAC and TRACERx cohorts. **b**. Time dependent ROC-AUC curves for CPTAC and TRACERx cohorts for 1,3,5-year intervals. **c**. For TRACERx cohort, comparing PATH-ORACLE to established risk factors for relapse in stage I LUAD. **d**. Subgroup analysis for pooled TRACERx and CPTADC cases. EGFR mutant cases are more accurately predicted. **e**. Correlation between of cases with preoperative ctDNA concentration and the PATH-ORACLE biomarker. **f**. TCGA-LUAD training data scaling ablation analysis. Model performance improves with training dataset size.

We compared risk stratification between ORACLE and PATH-ORACLE to assess the impact of the multimodal approach. In the TRACERx cohort (n=79), PATH-ORACLE corrected 19 ORACLE misclassifications (24.1%), reclassifying 5/17 relapse cases (29.4%) to high risk and 14/62 non-relapse cases (22.6%) to low risk. Accounting for remaining discordant classifications, this resulted in a net gain of 8 correctly stratified patients (+10.1%). In the CPTAC cohort (n = 68), PATH-ORACLE correctly reclassified 5 patients previously misclassified by ORACLE (7.4% of the cohort), including 1/13 relapse cases (7.7%) reassigned to high risk and 4/55 non-relapse cases (7.3%) reassigned to low risk. After accounting for cases in which PATH-ORACLE remained discordant with clinical outcome, this corresponded to a net improvement of 2 correctly stratified patients (+2.9%).

Motivated by prior studies reporting prognostic associations for molecular mutation markers (13,27), we assessed whether these variables were prognostic in our studied cohorts. In a mutation–survival analysis of 492 patients across four cohorts (TCGA-LUAD, TRACERx, CPTAC, and KORANYI), including 145 progression events, we found no statistically significant associations between EGFR, RAS, or ALK mutations and relapse (all p > 0.05). Although EGFR mutations exhibited a borderline protective trend (HR = 0.66 [0.41–1.05], p = 0.081), this effect was not reproducible across cohorts. These findings require validation in larger cohorts with more detailed molecular characterization, including variant allele frequency, specific mutation subtypes (for example, EGFR exon 19 deletions), and co-mutation patterns.

Despite having a poor prognostic power by themselves, we next evaluated whether these additional features provided a complementary prognostic signal in a multimodal setting. Mutation status, together with LVI, VPI, and clinical variables, were added to PATH-ORACLE, however, these additions did not consistently improve performance across validation cohorts. This indicates that the evaluated molecular and pathological variables do not add meaningful prognostic information beyond PATH-ORACLE.

We evaluated PATH-ORACLE’s discriminative ability (C-index with 95% CI via 1000 bootstrap iterations) across clinical and molecular subgroups (age, sex, EGFR/RAS/ALK mutation status, T-stage, and smoking status) in CRUK (n=79), CPTAC (n=68), and combined cohorts (n=147) **(Fig 3.d)**. In the combined cohort, PATH-ORACLE achieved an overall C-index of 0.747 (0.644-0.834) with consistent performance across most subgroups, but notably showed superior discrimination in EGFR mutant patients (C-index 0.811 [0.628-0.972], n=27, 5 events) versus EGFR wild-type (0.687 [0.539-0.829], n=85, 18 events), RAS wild-type patients (0.878 [0.787-0.951], n=58, 10 events) versus RAS mutant (0.559 [0.368-0.750], n=54, 13 events), and never smokers (0.915 [0.823-1.000], n=38, 3 events) versus ever smokers (0.792 [0.681-0.884], n=78, 20 events).

Preoperative ctDNA (6,28,29) and PATH-ORACLE scores were uncorrelated (R^2^=−0.01) (**Fig 3.e)** in the TRACERx cohort, suggesting that they may contain complementary information. Although there were only 12 patients with simultaneously available ctDNA, WSI and RNA-Seq data, the analysis suggests that ctDNA and the PATH-ORACLE biomarker capture distinct biological dimensions of risk.

Machine learning models often benefit from a larger amount of training data. To evaluate whether additional training data could improve PATH-ORACLE performance, we performed a data scaling ablation study on the TCGA-LUAD training cohort (n=274=100%) (**Fig 3 f.)** We systematically reduced the training set size by 10% of the original data. Random subsampling was repeated five times for each data scale to reduce variance in results. Model performance degraded smoothly as training data decreased. PATH-ORACLE improves by +0.014 C-index per 10% increase in training data, suggesting that scaling to larger datasets could further improve performance. This highlights the need for larger scale data collection for model training and external validation.

## 3 Discussion

The evaluation of H&E-stained whole slide images by pathologists has been an indispensable tool in the decision-making process for cancer therapy in patients. However, observer variability, especially when making quantitative estimates, introduces a significant uncertainty (30). Artificial Intelligence (AI)/Deep Neural Networks have provided a systematic, automated approach to extract robust features from pathological images allowing for more consistent evaluation (30–32). Recently published methods, such as ANORAK(9) and PATQUANT(32), use deep learning based segmentation for automated quantification of the previously identified morphological patterns commonly seen in LUAD. Using this approach we, similarly to previous studies, found a more consistent performance of predicting relapse of stage I LUAD over manual grading. These methods, however, were principally designed with the aim of performing a more robust reproduction of the morphological evaluation already established by human pathologists rather than discovering novel, so far hidden prognostic histological features.

Recent advances in self-supervised learning (33) have enabled WSI foundation models to encode histological slides as high dimensional feature vectors. The existence of “hidden” (spectrum of morphological signals, including subtle texture variations, spatial arrangements of cellular structures, and complex multi-scale patterns) features discovered by these foundation models is strongly suggested by several publications (34). For example, certain foundation model-extracted features were associated with EGFR and other gene mutations in LUAD (34), suggesting that the cancer driver mutation-associated biology may be reflected by thus far unidentified morphological features. Similar findings have been reported in prostate cancer, where deep learning models, using WSIs as input and trained directly on outcomes for biochemical recurrence have outperformed automated Gleason grading systems (10,11).

Based on this, we hypothesized that a similar strategy could identify fine-grained, relapse-specific morphological patterns in early-stage LUAD that may not have been previously described or codified in the IASLC grading system. We found that our deep learning-based predictor provided more accurate estimates of recurrence risk in stage I LUAD than AI-assisted IASLC grading. This improved performance may reflect its ability to integrate complex biological features. These features may include known associations between IASLC morphological subtypes and immune cell infiltration patterns. Prior studies have shown that low-risk tumors are characterized by low-grade lepidic growth and dense lymphocytic infiltration. In contrast, high-risk tumors exhibit solid or cribriform growth patterns, necrosis, and tissue discohesion with sparse lymphocytes (31). It may be clinically informative to reevaluate the large number of existing cohorts by these methods. These improved predictions may further aid therapeutic decision making in stage I cases.

Integrating diverse diagnostic modalities such as histology, molecular markers, and gene mutations has long posed a significant analytical challenge in cancer care. Clinicians must navigate fragmented data systems where information resides in separate databases with inconsistent formats, while manually extracting clinically relevant findings from lengthy technical reports under substantial time pressure. Modern machine learning allows for the integration of these complementary data modalities, enabling quantitative multimodal models that capture synergistic features associated with LUAD relapse.

There is increasing evidence that this approach, based on the combined evaluation of histology, gene expression, gene mutations etc., produces independently validated predictions that are more accurate than the predictions based on the various diagnostic modalities alone (35–38). Whole-slide image data captures tissue structure and cell morphology, while RNA-Seq captures changes in gene activity. Using both together helps explain ambiguous findings in either modality, improving prediction accuracy. To this end, PATH-ORACLE improves risk stratification by building on a prospectively validated biomarker (ORACLE). These results raise the possibility that other diagnostic features, such as radiology, time series of routine laboratory measurements etc. may further increase the accuracy of predicting recurrence in stage I LUAD.

The increased accuracy of predicting recurrence of stage I LUAD raises the question of how to incorporate this information into clinical therapeutic decision making. For stage IB tumors, due to the lack of consistent overall survival benefit in randomized trials (39), routine adjuvant platinum-based chemotherapy is not consistently recommended (3). On the other hand, the CALGB 9633 study reported survival benefit for stage IB tumors with a size ≥4 cm that were treated with adjuvant carboplatin—paclitaxel relative to non-adjuvant treated cases (40). Since worse survival in stage IB patients is mainly associated with relapse, it is likely that the observed survival benefit was at least partly due to delaying or preventing recurrence of disease. Therefore, prioritizing stage IB patients for adjuvant chemotherapy using an accurate method predicting relapse will likely improve overall survival sparing those patients of unnecessary chemotherapy side-effects that have low probability of recurrence. In fact, stage IB patients are offered adjuvant chemotherapy that are considered to be high risk based on histologic risk factors such as tumor size, tumor grade, lymphovascular invasion, and visceral pleural invasion (4). However, assessing recurrence risk based on these factors, that could be readily employed in the clinic, have not been unified in a consistent quantitative fashion, partly due to inconsistencies of detecting factors such as lymphovascular invasion (41). We were exploring here whether a deep learning method based on WSI and transcriptomics of surgical biopsies, that in principle could be processed in a centralized laboratory setting, could provide a more accurate and consistent alternative to histologic risk factors. The potential survival benefit of biomarker directed adjuvant chemotherapy in stage IB cases was recently highlighted by the HIGH-AIM trial (42).

In stage IA disease, all major guidelines (ESMO, ASCO, NCCN) recommend observation alone, as neither adjuvant chemotherapy, immunotherapy, nor targeted therapy has demonstrated a survival benefit in this population (39,43). However, high risk stage IA recurrence cases could be evaluated whether close monitoring by liquid biopsy based genetic analysis provides a robust early warning system. In case of a strong correlation, high relapse risk IA cases could be monitored in this manner and an earlier, more active therapeutic intervention could be evaluated for improving long term clinical outcome. This is especially relevant for EGFR-mutated lung adenocarcinoma where PATH-ORACLE is especially accurate. For stage IB cases adjuvant osimertinib is recommended based on the ADAURA trial (44) but the benefit of this treatment in stage IA is not known. Considering the relatively low recurrence rate in stage IA cases, establishing survival benefit of osimertinib treatment in a non-prioritized cohort may be more difficult and would require larger patient number than in a biomarker directed trial. Therefore, a reliable predictor of relapse may significantly improve treatment strategies in stage IA lung adenocarcinoma as well.

There are several limitations in this study to acknowledge. Firstly, our model is built only from untreated patients, which avoids confounding by indication and provides a clean estimate of *baseline* relapse risk. However, it cannot identify who would benefit from adjuvant therapy, since high untreated risk does not necessarily imply high treatment responsiveness. Interpretability of WSI-level foundation models remains limited: although prognostic signals for relapse can be captured, distilling these into human-interpretable, novel morphological insights beyond current knowledge is challenging; emerging vision–language models may partially mitigate this. Data availability is incomplete across cohorts (e.g., absence of RNA-seq for KORANYI), restricting validation of PATH-ORACLE. Furthermore, the numbers of cases are reccurances are low across validation cohorts. Foundation models do not fully correct for domain shift or scanner-related artifacts.

## 4 Methods

### 4.1 Patients and cohorts

This retrospective observational study involved the analysis of 710 patients’ data over four independent cohorts. Cases were chosen under a strict case selection process to examine tumor relapse biology as unambiguously as possible. For each cohort, we selected only pathological stage IA or IB cases (AJCC 9th edition (45)). According to the American Joint Committee on Cancer (AJCC) 9th Edition TNM classification, Stage IA and IB lung cancer (including lung adenocarcinoma) represent early-stage, localized disease without regional lymph node or distant metastasis (N0, M0). Stage IA includes tumors ≤ 3 cm in greatest dimension (T1a-T1c), confined to the lung or visceral pleura and without involvement of the main bronchus, whereas Stage IB comprises tumors **>** 3 cm to ≤ 4 cm (T2a) or tumors with limited local features such as visceral pleural invasion, involvement of the main bronchus (excluding the carina), or associated partial atelectasis or obstructive pneumonitis.

Some cohorts were collected during earlier editions of the AJCC staging system, but all cases were re-classified and re-graded by senior pathologists to align with the 9th edition Stage IA and IB criteria to ensure consistency across the study. All cases underwent lobectomy with no neo-adjuvant therapy received.

The primary endpoint studied was Relapse-Free Interval (RFI). RFI was strictly defined as the period of time following curative-intent surgical resection during which the patient remains alive and free of any evidence of disease recurrence. RFI begins on the date of definitive treatment (lobectomy) and ends at the first documented relapse, whether local, regional, or distant, confirmed by clinical, radiologic, or pathologic assessment. Patients who did not experience recurrence were censored at the date of last disease-free follow-up. The secondary endpoint was Overall Survival. This study analyzed four independent cohorts. CPTAC (USA) (n=71, 18.3% relapse events) had a median follow-up of 434 days, with Stage IA patients (n=27) showing 22.2% relapse at a median of 493 days and Stage IB patients (n=44) showing 15.9% relapse at a median of 911 days. KORANYI (Hungary) (n=249, 32.5% relapse events) demonstrated longer follow-up (median 1,673 days), with Stage IA patients (n=146, 30.1% relapse events) relapsing at a median of 814 days and Stage IB patients (n=103, 35.9% relapse events) at 724 days. TCGA-LUAD (USA) (n=275, 32.7% relapse events) had a median follow-up of 642 days and similar relapse rates between Stage IA (n=134, ∼29% relapse events) and Stage IB (n=136, ∼37% relapse events). Median time to relapse was 515 days for Stage IA and 524 days for Stage IB. TRACERx (UK) (n=115, 16.5% relapse events) showed a median follow-up of 1,136 days, with relapse occurring at a median of 537 days, and comparable relapse rates between Stage IA (n=66, 15.2% relapse events) and Stage IB (n=49, 18.4% relapse events).

#### 4.1.1 Cohorts overview

**Table 1.** Clinical cohorts for the study. These are LUAD Stage I cases that underwent Lobectomy.

| **Cohort** | **CPTAC (46)** | **KORANYI** | **TCGA-LUAD (47)** | **TRACERx (48)** |
| --- | --- | --- | --- | --- |
| Patients (clinical) | 71 | 249 | 275 | 115 |
| Stage IA (n, %) | 27 (38.0%) | 146 (58.6%) | 134 (48.7%) | 66 (57.4%) |
| Stage IB (n, %) | 44 (62.0%) | 103 (41.4%) | 136 (51.3%) | 49 (42.6%) |
| WSI slides | 172 | 293 | 272 | 120 |
| RNA samples | 231 | 0 | 293 | 190 |
| Patients with WSI | 68 | 249 | 272 | 112 |
| Patients with RNA | 71 | 0 | 275 | 81 |
| WSI + RNA patients | 68 | 0 | 272 | 79 |
| Age mean (range) (years) | 64.1 (40–78) | 63.45 (38–80) | 65.78 (33–88) | 69.9 (48–91) |
| Sex (M/F) | 45 / 26 | 75 / 174 | 115 / 160 | 42 / 73 |

All direct comparisons between modalities and models use the exact same subset of patients. Since not all cases have data for all modalities, each analysis uses a slightly different subset of cases. For example, the IASLC vs WSI foundation model comparison included patients with pathological grading and imaging data, while multimodal modeling (WSI, RNA, and WSI+RNA PATH-ORACLE) required patients with both imaging and transcriptomic data.

### 4.2 Derived histopathological features

ANORAK scores were directly calculated from WSIs using the official openly released pre-trained model (9) (rhttps://github.com/xi11/AIgrading). The implementation was run on the TRACERx H&E WSIs and compared to published values to ensure consistency. This pipeline was then applied to the remaining slides across the three other cohorts.

IASLC grades were provided for a subset of cases by the pathologists who reported on each cohort at the time of diagnosis. The IASLC was defined according to the updated definition (7).

#### 4.2.1 Whole slide image data acquisition and pre-processing

Formalin-fixed paraffin-embedded (FFPE) tissue sections (diagnostic slides) were collected for each case, stained with hematoxylin and eosin (H&E), and digitally scanned to generate whole slide images.

Whole slide images were collected for TCGA-LUAD (n=522, scanner=Aperio), CPTAC (n=172, scanner=Aperio), TRACERx (n=120, scanner=Hamamatsu), and KORANYI (n=296, scanner=3DHistech, Mirax). CPTAC and TCGA-LUAD slides were scanned at 20× magnification (mpp 0.49 µm and 0.26 µm), TRACERx at mixed 20×/40× (mean mpp 0.29 µm), and KORANYI at 20× (mpp 0.24 µm). Microns-per-pixel normalization was applied during preprocessing to ensure consistent spatial resolution across scanner platforms.

All slides were pre-processed by removing non-tissue background areas with the **Otsu** image processing method (49). Custom segmentation thresholds were chosen under visual examination for each cohort. RGB color-based exclusion was also applied to remove any markings on the slide. The segmented slides were then patched at 0.25 microns per pixel resolution (20X) using the Slide Crush package (https://github.com/ozkilim/SlideCrush). The resolution of each slide was determined from its metadata.

Each gigapixel whole slide image becomes a set of 256×256 pixel patches that form a tensor of RGB images shaped (n,3,256,256). These patches were embedded with the ConchV1.5 (17) patch-level foundation model to create a batch of embeddings shaped (n,783), where each patch becomes a single compressed vector describing the morphologies of that patch.

These embeddings were then passed through the pre-trained transformer model TITAN (18), which transforms the set of embeddings into a single embedding representing the entire slide. This pools the patch-level embeddings and compresses the information into one 783-dimensional vector. This vector is a dense representation of the slide’s local morphological patterns and global spatially resolved contextual relationships. This representation is then used for downstream learning.

#### 4.2.2 Whole slide image modeling

Models incorporating TITAN whole slide image embeddings were trained using TCGA-LUAD histopathology data and validated on the independent TRACERx, CPTAC, and KORANYI cohorts. Prior to modeling, the 783-dimensional TITAN embeddings underwent dimensionality reduction via principal component analysis (PCA) to compress the feature space while retaining the majority of variance to 24 dimensions. The TCGA-LUAD training cohort data was used to fit the transform, and the validation sets were transformed with the fixed PCA transform to avoid data leakage. Cox proportional hazards regression with elastic net regularization (α = 0.5, lambda optimized via 10-fold cross-validation) was employed for training to predict recurrence-free interval (RFI), incorporating both censored and non-censored patient outcomes as labels.

The Cox partial likelihood loss function (**Eq.1**) used for training is defined as:

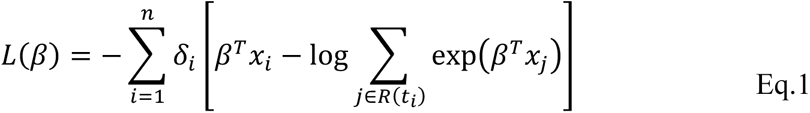

where **β** represents the coefficient vector to be estimated, **xi** is the covariate vector PCA-reduced TITAN embeddings for patient *i*, **δi** is the event indicator (1 if relapse occurred, 0 if censored) for patient *i*, **R(ti)** is the risk set at time *ti* containing all patients still at risk just before time *ti*, and **n** is the total number of patients. The elastic net regularization penalty was added to prevent overfitting:

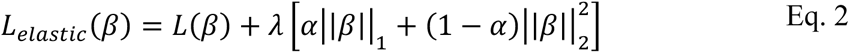

where **α = 0.5** balances L1 (lasso) and L2 (ridge) penalties, and **λ** was selected via 10-fold cross-validation to optimize predictive performance on the TCGA-LUAD validation sets.

For cases with multiple slides per patient, the fitted Cox model first generated a risk score prediction for each individual slide independently. These slide-level predictions were then mean-aggregated to produce a single patient-level risk score.

Kaplan-Meier survival curves and time-dependent ROC-AUC metrics were generated using the lifelines and scikit-learn Python packages, with survival probabilities estimated from Cox model risk scores and model discrimination evaluated at multiple time points. Hazard ratios stated in the text correspond to univariable Cox models comparing high- and low-risk groups.

### 4.3 RNA acquisition and processing

We harmonized published bulk RNA-seq data from CPTAC-LUAD GDC project CPTAC-3; dbGaP phs001287), TCGA-LUAD GDC project; dbGaP phs000178, and TRACERx EGA Dataset accession: EGAD00001009862 cohorts. A published set of poor-prognosis ‘Q4’ genes from the ORACLE study was used as the initial filter (11); we retained the Q4 genes present across all three cohorts, excluding any gene missing in a cohort and removing genes with very low total counts (TPM<10). Counts were coerced to non-negative integers, with duplicate sample identifiers disambiguated for multi-region profiles. Median-of-ratios normalization (DESeq2) (50) was applied, followed by a log1p variance-stabilizing transform. Batch effects were corrected with PyComBat (parametric prior, batch = cohort, no covariates) (51), and t-SNE embeddings were generated before/after batch correction for quality control (52). An ORACLE risk score was computed as a weighted linear combination of the 23 ORACLE signature genes using published coefficients (10).

We evaluated eight published prognostic gene signatures: ORACLE; Biswas Nat Cancer 2024 (11) (23 genes: ANLN, ASPM, CDCA4, ERRFI1, FURIN, GOLGA8A, ITGA6, JAG1, LRP12, MAFF, MRPS17, PLK1, PNP, PPP1R13L, PRKCA, PTTG1, PYGB, RPP25, SCPEP1, SLC46A3, SNX7, TPBG, XBP1), Carr J Thorac Cardiovasc Surg (20) (4 genes: CHEK1, KNSTRN, MIF, PAFAH1B3), Jin J Immunol Res 2022 (21) (5 genes: ADRB2, CCDC69, CCND2, IDH2, SFTPC), Li Sci Rep 2022 (22) (7 genes: CFTR, DDIT4, DERL3, IGFBP1, NUPR1, PDX1, PPP1R3G), Shang J Transl Med 2025 (23) (2 genes: COL11A1, THBS2), Song Sci Rep 2022 (24) (11 genes: ADM, GPC3, IL7R, NMI, NMUR1, PSEN1, PTPRE, PVR, SEMA4D, SERPINE1, SPHK1), Wang Front Immunol 2022 (25) (3 genes: HNRNPC, IGF2BP1, IGFBP3), and Zhao Lung Cancer 2020 (26) (19 genes: ABCC2, DKK1, EDN3, F2, FAM83A, GFRA1, GIP, IGF2BP1, IGFBP1, INHA, LINGO2, MS4A15, PEBP4, PSG1, RHCG, SH2D5, SPOCK1, UPK1B, ZIC5).

To identify the most promising signatures for this specific task, we first trained Cox proportional hazards models with elastic net regularization (α = 0.5, lambda optimized via 10-fold cross-validation) on TCGA-LUAD using each signature’s gene set independently, then evaluated each model’s generalization performance on the independent validation cohorts (TRACERx and CPTAC). The three signatures demonstrating the best generalization performance across validation cohorts were selected for further combined analysis and multimodal integration.

Models incorporating the full gene set from these top three performing signatures were retrained using TCGA-LUAD RNA-seq data with the same Cox elastic net approach (**Eq.1**) and validated on the independent TRACERx and CPTAC cohorts. For cases with multi-region RNA-seq samples, the fitted Cox model first generated a risk score prediction for each sample independently. These sample-level predictions were then mean-aggregated to produce a single patient-level risk score.

### 4.4 Mutation based analysis

Somatic mutation data for EGFR, KRAS/NRAS, and ALK were obtained from cohort-specific sources. These data were harmonized to form a binary mutation status. The harmonized data were used for downstream unimodal and multimodal analysis.

For CPTAC (n=33), gene-level mutation calls were extracted from the CPTAC whole-exome sequencing dataset (53), downloaded from the NCI Genomic Data Commons (GDC) Data Portal provided as a binary matrix with Ensembl gene identifiers indicating presence (1) or absence (0) of any somatic mutation per gene. For a subset of the KORANYI cohort (n=72), mutation status was derived from clinical molecular testing performed as part of routine diagnostic workup, with EGFR and KRAS mutations detected by real-time PCR and ALK rearrangements detected by fluorescence in situ hybridization or immunohistochemistry, recorded as binary values (0=negative, 1=positive) in institutional electronic medical records.

For TCGA (n=272), mutation calls were obtained from the MC3 (Multi-Center Mutation Calling in Multiple Cancers). Working Group gene-level mutation matrix for lung adenocarcinoma which aggregates somatic variants from whole-exome sequencing analyzed using multiple variant calling algorithms.

For TRACERx (n=115), mutation status was extracted from the curated annotation files containing binary calls (Yes/No) for each driver gene derived from the TRACERx 421 cohort’s targeted and whole-exome sequencing data (54).

Across all cohorts, samples were classified as mutant (MUT) if any pathogenic or likely pathogenic somatic alteration was detected in the respective gene, and wild-type (WT) if no pathogenic or likely pathogenic mutations were detected; for RAS status, samples were classified as RAS-mutant if either KRAS or NRAS harbored a mutation, and RAS wild-type if both genes were mutation-free.

Binary mutation status (mutant vs. wild-type) for EGFR, KRAS/NRAS, and ALK was used as input to Cox proportional hazards regression models with elastic net regularization to predict RFI.

Models were trained on TCGA-LUAD data and validated on the independent TRACERx, CPTAC, and KORANYI cohorts, consistent with the WSI and RNA-based analysis approach.

### 4.5 PATH-ORACLE - multimodal integration

#### Data harmonization and cohort overlap

Multimodal analysis was performed on patients having both WSI and RNA-seq data across three cohorts: TCGA-LUAD (n=272 patients with both modalities), CPTAC (n=68), and TRACERx (n=79). Only patients with complete data for the modalities being integrated were included in each analysis. For multi-slide or multi-region cases, modality-specific predictions were generated independently for each sample and then mean-aggregated to patient-level risk scores as described in the WSI and RNA modeling sections.

#### Feature preparation

Prior to integration, features from each modality were prepared as follows: WSI features: 24-dimensional PCA-reduced TITAN embeddings (described in WSI modeling section). RNA features: (46: genes from top 3 signatures) gene expression values, normalized and batch-corrected as described in the RNA processing section to ensure no leakage. Mutation features: 3 binary indicators (EGFR, KRAS/NRAS, ALK) for cases with available mutation data, clinical features: Age (continuous), sex (binary), stage (IA vs IB binary)]. All features were standardized scaled before concatenation to ensure comparable scales across modalities.

#### PATH-ORACLE - multimodal model architecture

We employed an early fusion strategy in which features from multiple modalities were concatenated into a single input vector prior to Cox regression. The multimodal risk model is defined as:

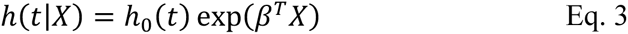

where **X** is the concatenated feature vector from the available modalities, **β** is the coefficient vector learned via elastic net-regularized Cox regression (consistent with unimodal models: α = 0.5, lambda optimized via 10-fold cross-validation on the training set), and **h₀(t)** is the baseline hazard function. Time-dependent ROC curves and AUCs were computed on the retained samples; 95% CIs came from 1,000 bootstrap resamples, and significance vs chance (AUC=0.5) used a 10,000-label permutation test.

#### Model configurations

We evaluated multiple model configurations to assess the incremental value of each added modality using only the TCGA-LUAD validation scores to guide model selection. Specifically, WSI only (n=272 TCGA, n=68 CPTAC, n=79 TRACERx), RNA only (n=272 TCGA, n=68 CPTAC, n=79 TRACERx), WSI + RNA (n=272 TCGA, n=68 CPTAC, n=79 TRACERx). All models were trained on the TCGA-LUAD training set and validated on independent TRACERx and CPTAC cohorts. The same elastic net Cox proportional hazards framework was used across all configurations to ensure fair comparison.

#### Comparison with circulating tumor DNA

We compared PATH-ORACLE model predictions with pre-operative circulating tumor DNA (ctDNA) measurements in a subset of TRACERx patients with available ctDNA data (n=12). ctDNA was quantified as tumor fraction in parts per million (ppm) using the tumor-informed **NeXT Personal** (**28**). This analysis assessed whether PATH-ORACLE and transcriptomic biomarkers provide complementary prognostic information to ctDNA-based minimal residual disease detection.

#### Training scale ablation

We subsampled the original training set of 274 cases at 50%, 60%, 70%, 80%, and 90% using 5 random seeds to create TCGA-LUAD training datasets of varying sizes. We retrained PATH-ORTACLE 5 times for each subset and averaged the C-index across runs for each training configuration. This provided a robust estimate of the data scaling laws for this task.

### 4.6 Compute and experimentation

All computational work was performed on the nemo HPC with nodes controlling A100 GPUs at the Francis Crick Institute, London.

## 5 Data availability

### 5.1 Ethics and institutional approvals

This study was conducted in accordance with the principles of the Declaration of Helsinki and approved by the relevant institutional review boards and ethics committees for each cohort.

TCGA-LUAD: Data were obtained from The Cancer Genome Atlas (TCGA) Research Network, a publicly available resource. All samples in TCGA were collected under protocols approved by local institutional review boards, with informed consent obtained from all participants. TCGA-LUAD whole slide images, RNA-seq data, and clinical annotations are publicly available through the Genomic Data Commons (GDC) portal (https://portal.gdc.cancer.gov/) and the Cancer Imaging Archive (TCIA, https://www.cancerimagingarchive.net/).

CPTAC: Clinical Proteomic Tumor Analysis Consortium data were accessed through the National Cancer Institute’s proteomic data portal. All original CPTAC sample collections were conducted under institutional review board approval with written informed consent from all participants. CPTAC proteomic, genomic, and imaging data are publicly available through the CPTAC Data Portal (https://proteomics.cancer.gov/data-portal) and the Cancer Data Service.

TRACERx: TRACERx (TRACERx 421) samples were collected as part of the TRACERx study under UK Research Ethics Committee approval. All participants provided written informed consent. Use of TRACERx data for this study was approved by the TRACERx Data Access Committee. TRACERx data are available through managed access via the European Genome-phenome Archive (EGA) upon application to the TRACERx Data Access Committee.

KORANYI: Retrospective analysis of clinical samples from the National Korányi Institute of Pulmonology, Budapest, Hungary, was approved by the Medical Research Council of Hungary (TUKEB) and the National Center for Public Health and Pharmaceuticals (NNGYK) (permit number: NNGYK/27869-5/2024). Patient consent was waived with the approval of the Medical Research Council since the study was conducted retrospectively on FFPE-archived resection samples.

#### Restricted access cohorts

Whole slide images and clinical information from the Korányi Institute cohort contain potentially identifiable patient information and are not publicly available due to ethical and legal restrictions. Access requests may be directed to the corresponding authors and will be considered on a case-by-case basis, subject to institutional review board approval and data use agreements. **Processed data:** Due to data use agreements and patient privacy considerations, processed feature matrices and model outputs cannot be made publicly available. Researchers may contact the corresponding author to discuss potential data sharing arrangements subject to appropriate institutional approvals.

## Data Availability

The datasets analyzed in this study are derived from a combination of publicly available and restricted-access sources. The Cancer Genome Atlas lung adenocarcinoma data, including whole slide images, RNA sequencing data, and clinical annotations, are publicly available from the Genomic Data Commons portal (https://portal.gdc.cancer.gov/) and The Cancer Imaging Archive (https://www.cancerimagingarchive.net/). Clinical Proteomic Tumor Analysis Consortium proteomic, genomic, and imaging data are publicly available through the Clinical Proteomic Tumor Analysis Consortium Data Portal (https://proteomics.cancer.gov/data-portal) and the Cancer Data Service. TRACERx data are available through managed access via the European Genome-phenome Archive, subject to approval by the TRACERx Data Access Committee. Whole slide images and clinical data from the National Koranyi Institute of Pulmonology cohort are not publicly available due to ethical and legal restrictions, as they contain potentially identifiable patient information. Access to these data may be granted upon reasonable request to the corresponding authors and subject to approval by the appropriate ethics oversight bodies and data use agreements. Processed feature matrices and model outputs generated in this study are not publicly available due to data use agreements and patient privacy considerations. Requests for access to these data may be directed to the corresponding authors and will be considered on a case-by-case basis.

## 6 Code availability

All analysis code, including preprocessing pipelines, model training scripts, and evaluation workflows, is publicly available at our GitHub repository: https://github.com/ozkilim/OncoOne. Pre-trained TITAN, ConchV1.5, and ANORAK models are available from their respective original publications. All code is provided under Apache 2.0. All analyses were conducted in Python 3.9.16 unless otherwise specified. Key software packages and their versions used in this study are listed below:

### Image processing and deep learning

PyTorch 2.0.1, torchvision 0.15.2, openslide-python 1.2.0, scikit-image 0.21.0, numpy 1.24.3, opencv-python 4.7.0, pillow 9.5.0

### Histopathology-specific tools

TITAN, ConchV1.5 ANORAK [https://github.com/xi11/AIgrading, https://zenodo.org/records/15272883] slide, crush trident

### RNA-seq processing and batch correction

DESeq2 1.38.3 (R 4.2.2), sva (ComBat) 3.46.0 (R 4.2.2), edgeR 3.40.2 (R 4.2.2), pandas 2.0.2, scanpy 1.9.3

### Survival analysis and machine learning

lifelines 0.27.7, scikit-survival 0.21.0, scikit-learn 1.3.0, glmnet (R implementation) 4.1-7 (R 4.2.2)

### Dimensionality reduction and visualization

scikit-learn (PCA, t-SNE) 1.3.0, matplotlib 3.7.1, seaborn 0.12.2, plotly 5.14.1

### Statistical analysis

scipy 1.10.1, statsmodels 0.14.0, rpy2 3.5.11 (for R interface)

### Reproducibility

Random seeds were set to ensure reproducibility where applicable (seed = 42 for Python random, numpy.random, and torch.manual_seed). Environment specifications and dependency management were handled using conda (version 23.3.1) with environment files provided in the code repository.

## ACKNOWLEDGMENTS

This work was supported by the Breast Cancer Research Foundation (BCRF-23-159 to Z.Szallasi), Kræftens Bekæmpelse (R325-A18809 and R342-A19788 to Z.S.), Det Frie Forskningsråd Sundhed og Sygdom (2034-00205B to Z.S.), NIH Grant 1 P01 CA228696-01A1 to Z.S and by the National Research, Development, and Innovation Office of Hungary via grant NKKP-153428 HIGHLIGHT (I,C.) and NKKP-152409 STARTING (O.P.). This work was also supported by the Ovarian Cancer Research Alliance by grant CRDGAI-2025-3-1992 to Z.S. and I.C. J.M. was supported by the Hungarian National Research, Development and Innovation Office (K147226). J.M., Z.Sz., and J.F. were supported by the Hungarian National Research, Development and Innovation Office (K147226). The results shown here are based upon data generated by the TCGA Research Network: http://cancergenome.nih.gov/ and the International Cancer Genome Consortium (ICGC): https://icgc.org/. C.S. is a Royal Society Napier Research Professor (RSRP\R\210001). His work is supported by the Francis Crick Institute that receives its core funding from Cancer Research UK (CC2041), the UK Medical Research Council (CC2041), and the Wellcome Trust (CC2041). For the purpose of Open Access, the author has applied a CC BY public copyright licence to any Author Accepted Manuscript version arising from this submission. C.S. is funded by Cancer Research UK (TRACERx (C11496/A17786), PEACE (C416/A21999) and CRUK Cancer Immunotherapy Catalyst Network); Cancer Research UK Lung Cancer Centre of Excellence (C11496/A30025); the Rosetrees Trust, Butterfield and Stoneygate Trusts; NovoNordisk Foundation (ID16584); Royal Society Professorship Enhancement Award (RP/EA/180007 & RF\ERE\231118); National Institute for Health Research (NIHR) University College London Hospitals Biomedical Research Centre; the Cancer Research UK-University College London Centre; Experimental Cancer Medicine Centre; the Breast Cancer Research Foundation (US) (BCRF-23-157); Cancer Research UK Early Detection an Diagnosis Primer Award (Grant EDDPMA-Nov21/100034); and The Mark Foundation for Cancer Research Aspire Award (Grant 21-029-ASP) and ASPIRE Phase II award (Grant 23-034-ASP). CS is in receipt of an ERC Advanced Grant (PROTEUS) from the European Research Council under the European Union’s Horizon 2020 research and innovation programme (grant agreement no. 835297). M.J-H has received funding from CRUK, NIH National Cancer Institute, IASLC International Lung Cancer Foundation, Lung Cancer Research Foundation, Rosetrees Trust, UKI NETs and NIHR. TRACERx is funded by Cancer Research UK (CRUK; no. C11496/A17786). We would like to thank Carlos Martinez Ruiz and Ariana Huebner for helping with oragniazation of data, computational resourses for experiements, and insights into the manuscript.

## Author contributions

O.K., Z.S., J.M., and C.S. conceived and designed the study. O.K. developed the methodology. O.K., J.M., J.F., C.N-L., S.V., and Z.S. acquired the data. O.K., Z.Szt., Z.S., O.P., J.M., D.A.M., M.D., A.H., M.J-H., and A.P. analyzed and interpreted the data (Analysis and interpretation of data (e.g., statistical analysis, biostatistics, computational analysis)). Z.S., I.C., and C.S. supervised the study.

All authors contributed to the preparation of the manuscript and the Supplementary Materials.

---

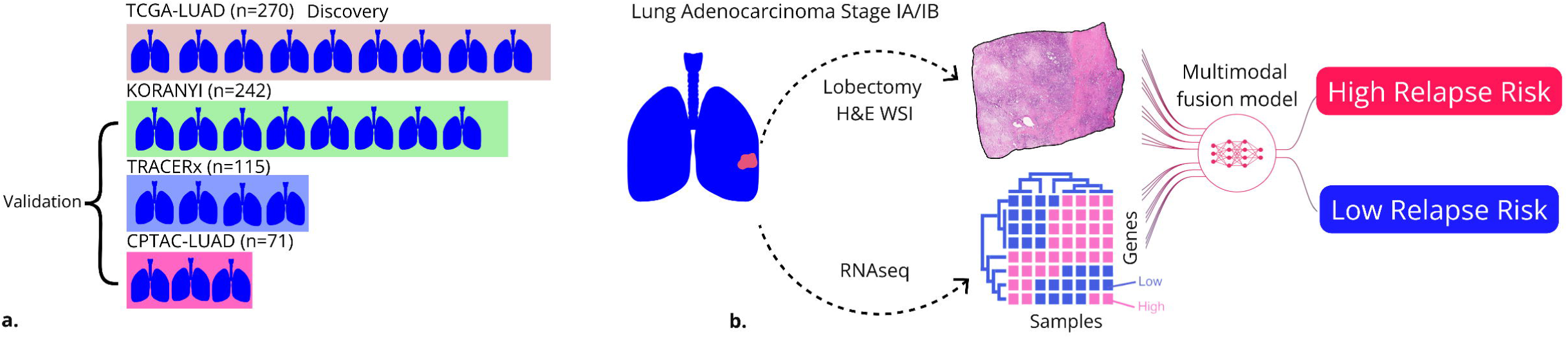

